# Characterizing Spatial Epidemiology in a Heterogeneous Transmission Landscape Using a Novel Spatial Transmission Count Statistic

**DOI:** 10.1101/2023.12.28.23300535

**Authors:** Leke Lyu, Gabriella Veytsel, Guppy Stott, Spencer Fox, Cody Dailey, Lambodhar Damodaran, Kayo Fujimoto, Pamela Brown, Roger Sealy, Armand Brown, Magdy Alabady, Justin Bahl

## Abstract

**Background:** Viral genomes contain records of geographic movements and cross-scale transmission dynamics. However, the impact of regional heterogeneity, particularly among rural and urban centers, on viral spread and epidemic trajectory has been less explored due to limited data availability. Intensive and widespread efforts to collect and sequence SARS-CoV-2 viral samples have enabled the development of comparative genomic approaches to reconstruct spatial transmission history and understand viral transmission across different scales.

**Methods:** We proposed a novel spatial transmission count statistic that efficiently summarizes the geographic transmission patterns imprinted in viral phylogenies. Guided by a time-scaled tree with ancestral trait states, we identified spatial transmission linkages and categorize them as imports, local transmissions, and exports. These linkages were then summarized to represent the epidemic profile of the focal area.

**Results:** We demonstrated the utility of this approach for near real-time outbreak analysis using over 12,000 full genomes and linked epidemiological data to investigate the spread of the SARS-CoV-2 in Texas. Our study showed (1) highly populated urban centers were the main sources of the epidemic in Texas; (2) the outbreaks in urban centers were connected to the global epidemic; and (3) outbreaks in urban centers were locally maintained, while epidemics in rural areas were driven by repeated introductions.

**Conclusions:** In this study, we introduce the Source Sink Score, which allows us to determine whether a localized outbreak may be the source or sink to other regions, and the Local Import Score, which assesses whether the outbreak has transitioned to local transmission rather than being maintained by continued introductions. These epidemiological statistics provide actionable information for developing public health interventions tailored to the needs of affected areas.

**Plain Language Summary:** This study examined how COVID-19 spread through urban and rural areas in Texas by analyzing the virus’s genomes from over 12,000 samples. Our goal was to understand how the virus travels and impacts different regions. Our findings revealed that densely populated urban centers were the primary sources of the virus in Texas. In contrast, outbreaks in rural areas were often fueled by new introductions of the virus from external sources. To conduct this analysis, we employed new computational methods that track where the virus originates and where it spreads. These methods provide detailed information crucial for public health officials, particularly in regions where the virus has more severe impacts or exhibits unique spread patterns.

## Introduction

Genomic epidemiology is a field that utilizes pathogen genomes to study the spread of infectious diseases through populations ^1^. This approach has become increasingly popular due to the decreasing cost of genomic sequencing combined with increasing computational power. During the COVID-19 pandemic, increased number of countries started generating genomic data to inform public health responses ^2^. The Global Initiative on Sharing All Influenza Data (GISAID) ^3^ expanded to accommodate these novel data and now maintains the world’s largest database of SARS-CoV-2 sequences. As of December 2023, over 16 million sequences, sampled from over 200 countries/regions, have been submitted and archived. Such a vast and diverse dataset enables researchers and public health officials to identify key mutations ^4,5^ and track the emergence of variants of interest (VOIs) or variants of concern (VOCs). Additionally, this wealth of genomic information creates opportunities to uncover the hidden characteristics of the local-scale outbreak, such as the spatial dispersal of transmission and the demographic characteristics contributing to transmission patterns. However, effectively handling the complexity of the SARS-CoV-2 genomic dataset requires addressing key challenges, such as establishing robust sampling frameworks to draw reliable conclusions and developing efficient computational algorithms/pipelines.

In genomic epidemiology, analyzing sampling biases and develop an appropriate sampling strategy are crucial steps ^6^. Recent studies have shown that differences in epidemiology and sampling can impact our ability to identify genomic clusters ^7^. For instance, decreased sampling fraction can lead to the identification of multiple, separate clusters. Sampling biases can also impact phylogeographic analyses. When investigating diffusion in discrete spaces, if a specific area is overrepresented in the dataset, it may lead to an overrepresentation of the same area at inferred internal nodes ^1^. Similarly, when investigating diffusion in continuous space, extreme sampling bias might cause the posterior distribution to exclude the true origin location of the root ^8^.

Viral transmission happens at different spatial scales, encompassing international pandemics, domestic dispersal, and local outbreaks such as those in jails, nursing homes, hospitals, or schools. By mapping how pathogens spread through space and time, evidence-based interventions can be better developed and applied across various scales ^9^. The well-established software package, Bayesian Evolutionary Analysis Sampling Trees (BEAST) ^10^, implements discrete ^11^ and continuous^12^ phylogeographic models. Previous studies have used the discrete model to identify the transmission clusters of SARS-CoV-2 introduced in Europe ^13^, United States ^14^, Denmark ^15^ and England ^16^. Additionally, the continuous model has been applied to elucidate the spatial expansion of SARS-CoV-2 in Belgium ^17^ and New York City ^18^. Moreover, the BEAST module can accommodate individual travel history ^19^ to yield high-accuracy prediction regarding the location of ancestral nodes. Apart from Bayesian analysis, TreeTime ^20^ applies a maximum likelihood approach to infer the transitions between discrete characters. As a component of the Nextstrain ^21^ pipeline, this fast analysis enables real-time tracking of pathogens. With the rapid growth in SARS-CoV-2 genome data, we are now facing extensive phylogenies with thousands of tips. This raises the question: How can we translate the evolutionary changes of geographic traits from such expansive trees into clear epidemiological insights?

The transmission dynamics of SARS-CoV-2 are shaped by host immunity, host movement patterns, and other demographic characteristics ^22^. For instance, in Chile, people aged under 40 in municipalities with the lowest socioeconomic status had an infection fatality rate 3.1 times higher than those with the highest socioeconomic status ^23^. The severity of SARS-CoV-2 infection and the risk of mortality increased significantly with age ^24^. Accordingly, identifying at-risk populations is crucial for determining the potential burden on public health. In the US, rural populations have been particularly vulnerable to COVID-19 complications ^25^, experiencing higher incidences of disease and mortality ^26^. This vulnerability is largely attributed to limited access to healthcare and social services ^27^, as well as reduced access to and utilization of health information sources ^28^ compared to urban residents. Previous phylodynamic analyses have shown that frequent bi-directional transmission occurs between rural and urban communities ^29^. However, few studies have investigated the differences in transmission patterns between these areas.

In this study, we developed a pipeline to understand local-scale epidemic trends. Our approach includes proportional genome sampling based on case counts ^14^, followed by phylogeographic analysis using the Nextstrain framework ^21^. Lastly, we summarize and compare transmission patterns across subregions to identify viral sources and sinks. To demonstrate the utility of this method, we focused on the Delta wave in Texas, aiming to characterize viral diffusion within the state and compare epidemic trends between urban and rural areas.

## Methods

### Surveillance and genetic dataset

The United States Office of Management and Budget (OMB) defines Texas as having 25 metropolitan areas (Table S1). Any population, housing, or territory not included in these metropolitan areas is classified as rural. The Rural-Urban Continuum Codes (RUCC) further categorize metropolitan areas based on population size. Dallas–Fort Worth, Houston, San Antonio, and Austin, all classified as RUCC-1 ^30^, represent the most populous metropolitan areas in Texas.

We obtained historical COVID-19 data for confirmed cases in Texas from the Texas Department of State Health Services (DSHS) website ^31^. These weekly case counts, organized by county, were aggregated into metropolitan areas to inform our genome sampling strategy. Following the approach of Anderson F. Brito ^14^, we developed R scripts, later consolidated into an R package called *Subsamplerr*. This package processes case count tables and genome metadata, enabling visual exploration of sampling heterogeneity and the implementation of proportional sampling schemes.

With support from the Houston Health Department (HHD), we accessed a large dataset of SARS-CoV-2 genomes sampled in Texas: 51,229 genomes with linked metadata. We focused on the Delta variant for our analysis, as its outbreak caused severe illness, spread rapidly before widespread immunity was established, and was intensely sampled at multiple scales ^32^. Of the available genomes, 24,593 were identified as Delta variant, and 5,899 were subsampled proportionally to the case counts. Additionally, we sampled 6,386 Delta genomes from 49 countries to provide global context and estimate viral migration to and from Texas. Our final dataset comprises 12,285 epidemiologically linked SARS-CoV-2 genomes.

### Phylogeographic analysis pipeline

The pipeline comprises two major components: (1) Phylogenetic Reconstruction and (2) Characterization of Spatial Transmission Linkages.

Phylogenetic Reconstruction: This component utilizes the Nextstrain pipeline ^21^ to generate a time-labeled phylogeny with inferred ancestral trait states. Sequence alignment was conducted using Nextalign ^21^, while the maximum likelihood tree construction was achieved with IQ-TREE ^33^, applying a GTR substitution model. TreeTime ^20^ was employed to produce a time-scaled phylogeny and infer ancestral node states. The phylogeny was rooted using early samples from Wuhan (Wuhan-Hu-1/2019). Its temporal resolution was set based on an assumed nucleotide substitution rate of 8 ∗ 10^-4^ substitutions per site per year (default setting of Nextstrain build for SARS-CoV-2). Migration patterns between distinct geographic regions were inferred through time-reversible models ^20^.

Characterization of Spatial Transmission Linkages: This component used custom scripts to identify spatial transmission linkages from the phylogeny and summarize epidemic trends in the focal region. The tree file was imported and read using the ‘treeio’ ^34^ package in R. The tree was then converted into a structured data frame for further analysis, facilitated by the ‘tidytree’ package ^34^. Branches with durations exceeding 15 days were excluded, and the shorter branches in the phylogeny were designated as spatial transmission linkages. By analyzing trait states, we identified whether transmissions occurred within the focal area, involved imports from another location, or resulted in exports to another area. The time series of spatial transmission counts, categorized by type, provides an overview of the epidemic trends in the focal region.

### Metrics that describe transmission pattern

Different areas possess varying population sizes, levels of population mobility, and immunological characteristics, all of which can contribute to differences in the size and dynamics of the epidemic. We introduced two metrics to quantitatively compare the characteristics of epidemics in different areas.

We define the Local Import Score to estimate the proportion of new cases due to introductions:

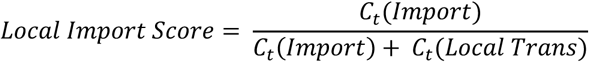

*C*_*t*_(*Import*) represents the count of viral imports over a specific period *t* and *C*_*t*_(*Local Trans*) represents the count of local transmissions during the same period. The choice of the time window for calculation is contingent on the research objective. It can encompass the entire duration of the epidemic wave to assess cumulative effects, or it might focus on shorter intervals, such as epidemiological weeks, for real-time surveillance. The Local Import Score ranges between 0 and 1.

We introduce the Source Sink Score to identify whether a region acts primarily as a viral source or sink:

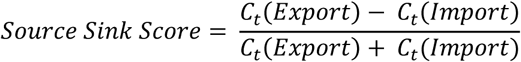

*C*_*t*_(*Export*) represents the count of exports over a specific period *t*. The Source-Sink Score ranges from -1 to 1. A score close to 1 indicates that the region primarily acts as a viral source, while a score near -1 suggests that the region mainly functions as a viral sink.

### Phylogenetic-based spatial network

We constructed a weighted, undirected network to capture the viral flow between metropolitan areas in Texas. Each metropolitan area is represented as a node, and the edge carries weight corresponding to the spatial transmission counts. After establishing the network, we conducted the centrality analysis to rank the metropolitan areas based on their betweenness, closeness, and degree centrality. We processed the various network data objects using the ‘igraph’ package ^35^ in R. Visualizations were generated with the ‘ggplot2’ package ^36^. We utilized the ‘qgraph’ package ^37^ to compute the centrality statistics of nodes.

### Sensitivity analysis of Source Sink Score and Local Import Scores

To evaluate the robustness of the Source Sink Score and Local Import Score, we conducted a sensitivity analysis by generating nine additional genome datasets for Texas. These datasets were created using the same proportional sampling scheme as the original dataset. We then ran the same phylogeographic workflows on each dataset. By comparing the results across these replicates, we assessed how uncertainties in sampling and phylogenetic inference affected the calculated scores.

## Results

### Genome sampling bias and subsampling scheme adjustments

With support from the Houston Health Department (HHD), we collected 24,593 Delta samples (B.1.167.2 and AY*) with high-coverage complete genomes (>29,000 bp) and linked sampling site ZIP codes. Our genome database contained over a thousand distinct ZIP code records, which we translated into their affiliated metropolitan areas. We calculated the sampling ratio by dividing the number of available genomes by the number of reported cases to explore sampling biases. Significant heterogeneity in sampling ratios was observed across different metropolitan areas from Epi-Week 14 to Epi-Week 43 (Figure S1A). Victoria, Wichita Falls, and Bryan-College Station were identified as the top three under-sampled metropolitan areas, while Houston, San Angelo, and Abilene were the most over-sampled. To mitigate potential sampling biases, we applied a proportional sampling scheme (Figure S1B), thereby enhancing the accuracy of our phylogeographic analysis ^9,10^. We adopted a consistent sampling ratio of 0.006 as a baseline for all regions. In regions that were under-sampled (sampling ratio below the baseline), all available genomes were retained. Conversely, over-sampled regions (with a sampling ratio exceeding the baseline) were down-sampled to match the baseline rate. As a result, we selected 5,899 Texas genomes, and the variance in sampling ratios across all metropolitan areas dropped substantially from 5.74e-05 to 7.56e-07.

### The transmission dynamics in Texas

We conducted a comprehensive phylogeographic analysis of 12,048 SARS-CoV-2 Delta genomes sampled from March 27, 2021, to October 24, 2021, to investigate the timing of virus introduction into Texas and the dynamics of the resulting local transmission lineages. These genomes were selected to ensure a roughly 1:1 ratio between Texas sequences (Table S2) and globally contextual sequences (Table S3). The Nextstrain ^21^ phylogenetic workflow was applied, in which a phylogenetic tree was estimated using IQ-TREE ^33^, and a time-adjusted phylogeny was inferred with TreeTime ^20^. The trait states of ancestral nodes were reconstructed as either ‘Texas’ or ‘non-Texas’ using the ‘mugration’ model implemented in TreeTime.

By considering the branches connecting each node to its parent as spatial transmission links, the location trait assigned to the nodes helps us categorize these connections into imports, local transmissions, and exports (Figures 1A, 1C). We defined a time series for these links as spatial transmission counts, providing a comprehensive summary of the epidemic’s trends over time (Figures 1B, 1D). Given that the infectious period for SARS-CoV-2 typically ranges from day 2 to day 15 post-infection ^22^, longer branches in the phylogeny likely indicate multiple transmission events. To reduce uncertainty, we excluded branches with durations exceeding 15 days, removing 9,995 out of 22,991 branches. Our findings reveal that the Delta variant was first introduced into Texas on April 5, 2021, with a confidence interval from March 18, 2021, to April 5, 2021, preceding the first documented case in Houston in mid-April 2021 ^39^. The Texas epidemic featured at least 311 viral imports and 433 viral exports, linking statewide cases to the global pandemic. The outbreak in Texas was predominantly driven by local transmission, with 6,584 branches classified as local transmission.

**Figure 1.**
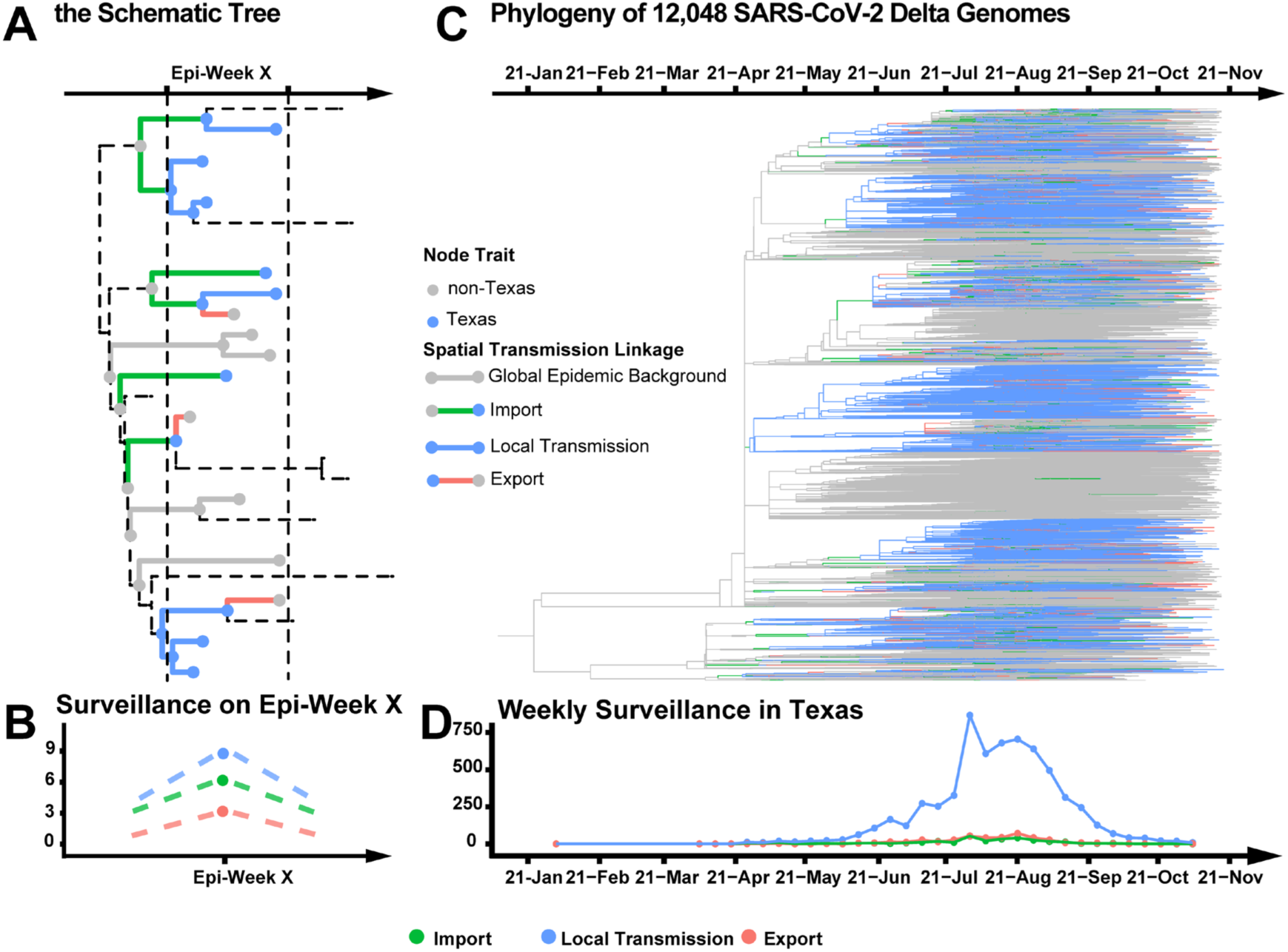
The spatial transmission count statistic investigates the transmission dynamic. **A.** Conceptual figure showing that transmissions can be classified into three categories: import, local transmission, and export. **B.** The schematic tree depicts a total of 18 spatial transmission linkages in Epi-Week X: 6 imports, 9 local transmissions, and 3 exports. **C.** In the time-adjusted phylogeny, branches are colored based on the categories of the corresponding spatial transmission linkages. **D.** The time series of spatial transmission counts summarizes the epidemic trend in Texas.

### Characterizing spatial transmission heterogeneity

To understand the spatial transmission of SARS-CoV-2 in Texas, we estimated ancestral location states on the phylogeny described above, incorporating 27 location traits: one contextual trait and 26 subregions of Texas (25 metropolitan areas and one combined rural area) (Figure S2). We then constructed a network of metropolitan areas in Texas based on phylogeographic signals (Figure 2A). The inferred network consisted of 25 nodes and 88 edges. Centrality analysis, detailed in Table S4, highlighted four pivotal nodes: Dallas–Fort Worth, Houston, San Antonio, and Austin.

**Figure 2.**
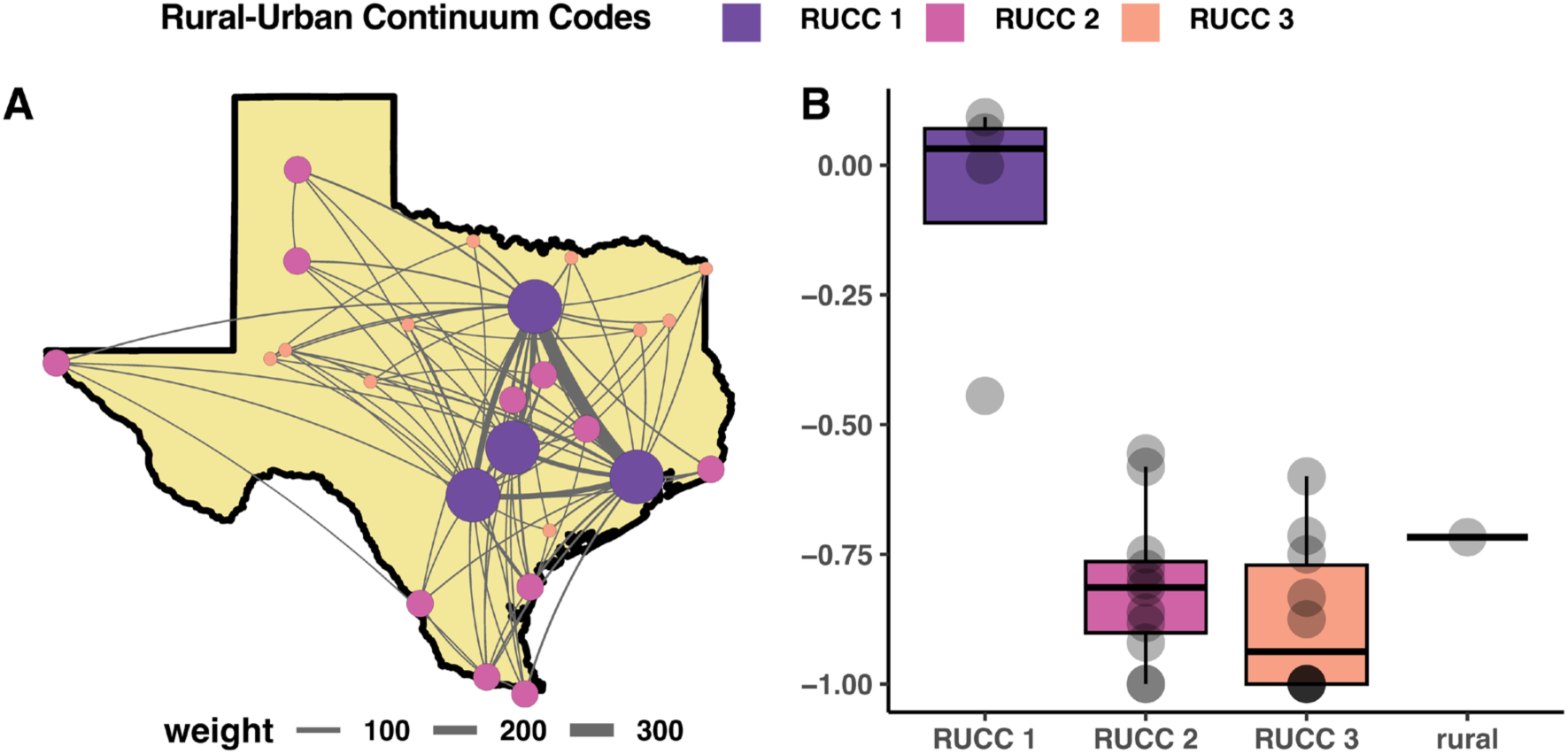
Characterizing Spatial Transmission Heterogeneity. Subregions across Texas are categorized by their Rural-Urban Continuum Codes (RUCC). RUCC-1 includes metropolitan areas with over 1 million residents, RUCC-2 includes areas with populations between 250,000 and 1 million, and RUCC-3 represents areas with fewer than 250,000 residents. The four major urban centers—Dallas-Fort Worth, Houston, San Antonio, and Austin—are classified as RUCC-1. **A.** Phylogeographic network of Texas metropolitan areas. In this network, each node represents a metropolitan area, and the width of the edges is proportional to the spatial transmission counts. **B.** The Source Sink Score identifies key source hubs of SARS-CoV-2 spread in Texas. Dots in the box plot represent subregions of Texas.

These subregions were consistently identified as key hubs based on degree, betweenness, and connectedness ^40^. Notably, all four of these metropolitan areas are classified as RUCC-1, suggesting populated urban centers played a crucial role in the viral spread across Texas.

### Community source-sink dynamics

We introduced the Source Sink Score to classify populations as either viral sources or sinks. This score ranges from -1 to 1, with a score near 1 indicating a population is predominantly a viral source—where the number of exports greatly exceeds imports—and a score near -1 indicating a population is primarily a viral sink, where imports dominate over exports.

Subregions of Texas were categorized as sources or sinks based on their cumulative Source Sink Score, with the full list provided in Table S5. Our analysis showed that the RUCC-1 group, which represents densely populated urban centers, had the highest Source Sink Scores, emphasizing its role as a major source during the outbreak in Texas (Figure 2B). Within the RUCC-1 group, Dallas-Fort Worth had the highest score at 0.092, followed by Houston (0.063), San Antonio (0.000), and Austin (-0.444). In contrast, rural areas, with a score of -0.717, primarily acted as viral sinks.

### Epidemic trends in populated urban centers compared to rural areas

We introduced the Local Import Score to estimate the proportion of new cases due to introductions. This score ranges from 0 to 1, with values closer to 1 indicating that the outbreak is primarily driven by external introductions, and values closer to 0 suggesting that local transmission is well-sustained. Identifying when most new cases are locally acquired is crucial for informing public health resource allocation, contact tracing efforts, and control strategies during emergency situations.

Using Houston as a representative city, we compared epidemic trends in densely populated urban centers to those in rural areas (Figure 3). Epidemic trends for other subregions are shown in Figures S3–S26. The accumulated Local Import Score for Houston during the entire Delta wave was 0.176, indicating that the outbreak was largely sustained by local transmission. In contrast, rural areas had an accumulated Local Import Score of 0.558, suggesting that the epidemic there was primarily driven by external introductions. Our results suggest that while an outbreak may initially rely on external introductions, once the epidemic becomes locally sustained, the region can evolve into a primary source of pathogen spread to other areas (Figures 3C and 3D).

**Figure 3.**
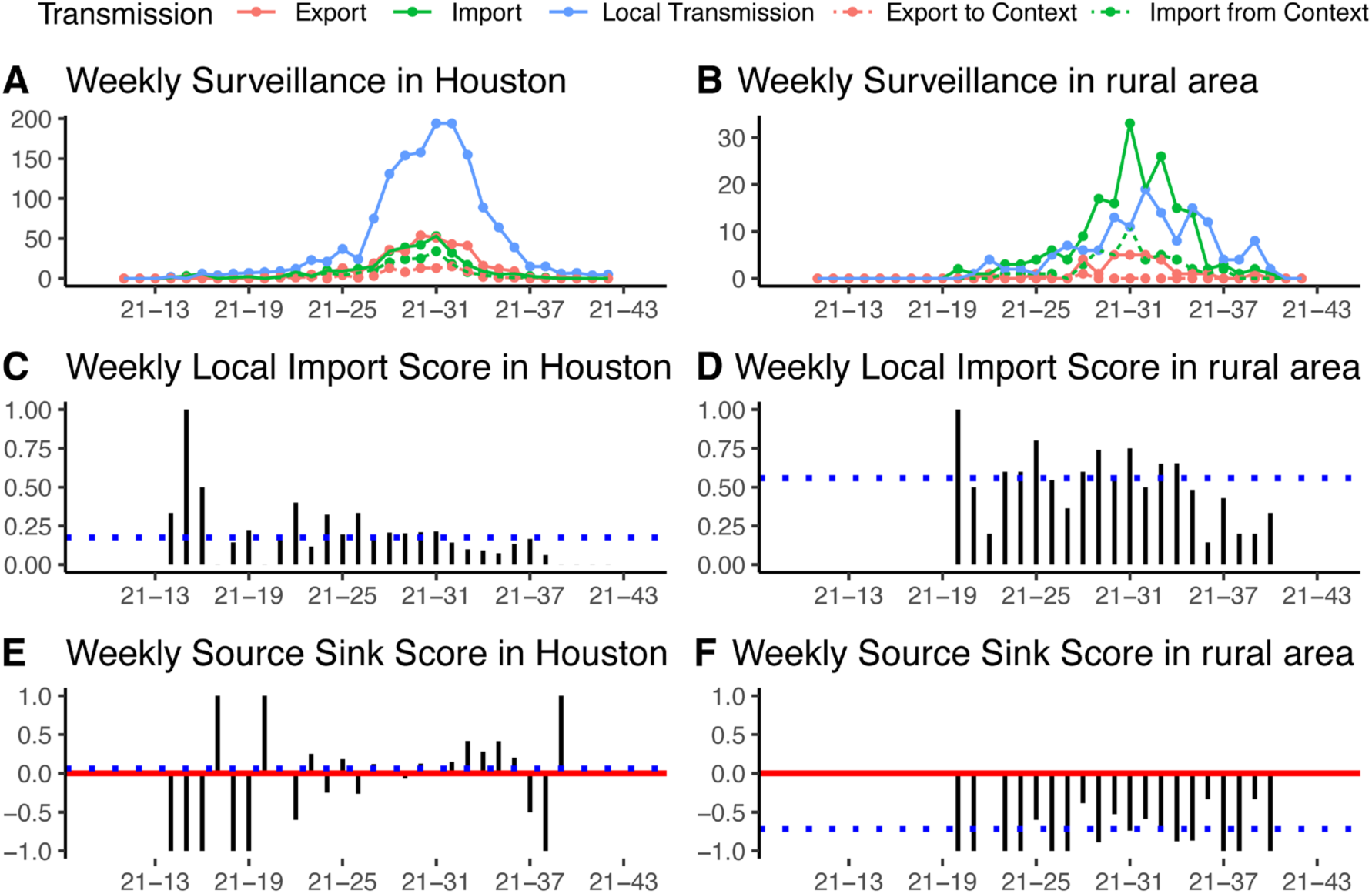
Epidemic trends on the Delta outbreak in populated urban centers vs the rural areas. **A.** The epidemic trend of Houston. **B.** The epidemic trend of the rural areas. The top of the panel shows the time series of spatial transmission counts by week. The dashed pink line represents exports from the analyzed regions to non-Texas. The dashed green line represents imports from non-Texas into the analyzed regions. **C.** The trend of Local Import Score in Houston. **D.** The trend of Local Import Score in rural areas. The black bars in the middle of the panel depict the weekly dynamics of Local Import Score. The dashed blue line indicates the accumulated Local Import Score. **E.** The trend of Source Sink Score in Houston. **F**. The trend of Source Sink Score in rural areas. The solid red line represents the benchmark of 0, indicating a balance between imports and exports. The dashed blue line marks the accumulated Source Sink Score.

We also analyzed viral flow between global contexts and urban centers (e.g., Houston) (Figure 4A), as well as between global contexts and rural areas (Figure 4B). Introductions from outside Texas accounted for 60% of all imports to Houston, while 25% of all exports from Houston were to locations outside Texas. By comparison, introductions from non-Texas sources accounted for 26% of all imports to rural areas, and 3% of rural exports were to locations outside Texas. These findings suggest that Houston, as a highly connected and large urban center, served as an important hub linking the outbreak across Texas to the broader global pandemic.

**Figure 4.**
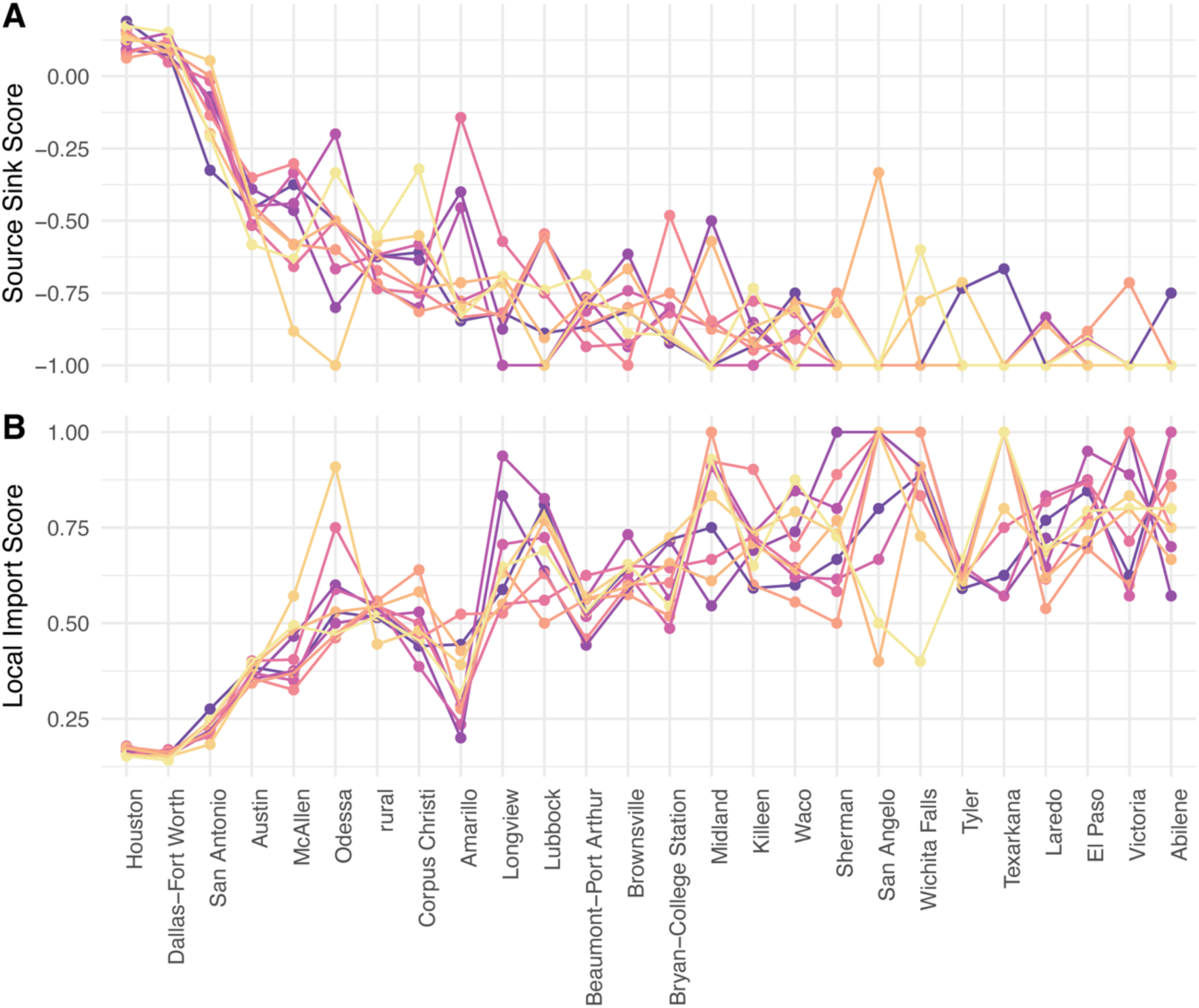
Sensitivity Analysis of 10 Replicates. **A.** Source Sink Score for subregions in Texas. **B.** Local Import Score for subregions in Texas. Both plots share the same x-axis, where regions are ranked from highest to lowest mean Source Sink Score. Each colored line connects the statistics estimated from the same replicate.

### Assessing the sensitivity of the new metrics

Despite the uncertainties inherent in sampling and phylogenetic reconstruction, our previous conclusions remained consistent across replicates. All 10 replicates supported RUCC-1 regions as the predominant viral sources, as these regions consistently showed the highest Source Sink Scores (Figure 4). Houston and Dallas–Fort Worth displayed the most robust results, as reflected by their narrow score ranges. Specifically, the Source Sink Score for Dallas–Fort Worth ranged from 0.049 to 0.151, while Houston’s score ranged from 0.063 to 0.190. The Local Import Score for Dallas–Fort Worth ranged from 0.142 to 0.169, while Houston’s score ranged from 0.152 to 0.178. A detailed record of the sensitivity analysis conducted across different metrics is provided in Table S6.

## Discussion

In this study, we introduced a novel spatial transmission count statistic, which characterizes the weekly counts of local spread, viral inflow, and outflow, illustrating transmission trends over time. The Source Sink Score and Local Import Score are heuristic metrics that allow for quantitative comparison of epidemic trends between regions. The Source Sink Score measures net viral exports, weighted by the outbreak size, while the Local Import Score compares the significance of external introductions versus local transmission in shaping the epidemic. We investigated the geographic diffusion pattern of SARS-CoV-2 in Texas to demonstrate the utility of this novel phylogeographic approach. At the state level, we characterized the timing and size of viral imports. Within the state of Texas, we reconstructed regional dissemination and contrasted the epidemic trends between urban centers and rural areas.

The size of our genomic data offers unprecedented opportunities for high-resolution investigations of spatial transmission history. Our analysis revealed that cryptic transmissions began as early as late March, 2 to 3 weeks before the identification of the first Delta case in Houston ^39^. Additionally, we identified at least 311 imports and 433 exports, highlighting Texas’s intensive connection to the global pandemic. Our results indicated that the Delta variant invaded Texas through multiple introductions. These independent imports subsequently formed massive local transmission clusters in Texas. This pattern aligns with observations from Connecticut’s initial COVID-19 wave ^41^, the UK’s first wave ^42^, the emergence of B.1.1.7 variant across the United States ^14^, and the presence of Omicron BA.1 in England ^16^.

The spatial transmission count statistic represents the time-series of categorized transmission linkages related to the focal regions. Informed by the annotated viral phylogeny, it summarizes the trends of local spread and viral flow at a minimal computational cost. Adopting a simplified model, we assume that transmission events take place along all the branches of the viral phylogeny. However, phylogenetic trees are not equivalent to transmission trees; they do not directly reveal who infected whom ^43,44^. As a result, our model may introduce bias in the estimation of local transmission counts. Despite this limitation, it provides valuable insights into local-scale transmission and epidemic trajectories that can inform control efforts. The efficiency of this statistic enables real-time surveillance of tens of thousands of viral genomes, which is crucial for addressing the challenges posed by the current pandemic or potential future outbreaks.

The role of a population as a source or sink evolves dynamically as the outbreak progresses and host immunity develops. Therefore, the Source Sink Score should be interpreted as a comparative measure, emphasizing relative differences between regions rather than absolute values. In Texas, populated urban centers functioned as the primary viral sources during the outbreak. Among all subregions, the RUCC-1 group had the highest Source Sink Scores, with Dallas-Fort Worth had the score at 0.092, followed by Houston (0.063), San Antonio (0.000), and Austin (-0.444). The significant role of these urban centers in spreading the epidemic can be linked to their key locations in road and air travel networks. Houston, Dallas-Fort Worth, and San Antonio, connected by Interstates 10, 45, and 35, form the vertices of the Texas Triangle ^45^, one of 11 megaregions in the US and home to the majority of the Texas’s population. This complex connectivity, along with the presence of major airports such as George Bush Intercontinental Airport in Houston (a United Airlines hub), Dallas-Fort Worth International Airport (American Airlines’ largest primary hub and headquarters), and San Antonio International Airport (a Southwest Airlines hub), highlights their pivotal role in airway travel. Our analysis underscored the crucial role of urban centers in driving the outbreak. This insight provides valuable information that can guide public health decision-making. Increased control efforts in highly connected urban centers may have a disproportionate impact on connected rural areas ^46^.

Rural areas exhibit a lower level of viral flow in relation to global contexts, with epidemics in these regions predominantly relying on external introductions, thus establishing them as viral sinks. Notably, urban centers and rural areas demonstrate distinct transmission patterns. It is important to note that our analysis assumes that virus transmission in each region is influenced only by population size and density, without accounting for the effects of community behavior and beliefs, healthcare disparities, environmental factors, and other influences on viral transmission. Future studies addressing these aspects will provide more comprehensive insights into the underlying drivers of transmission.

Despite uncertainties in sampling and phylogenetic reconstruction, all replicates from the sensitivity analysis supported RUCC-1 regions as the predominant viral sources. The robustness of both the Source Sink Score and the Local Import Score varied across regions. Houston and Dallas–Fort Worth exhibited more stable results, with narrower score ranges, likely due to the larger volume of data available (>1500 genomes). In contrast, regions such as Amarillo, Odessa, and San Angelo had fewer genomes (<50 genomes), leading to broader score ranges and making interpretation less reliable. We believe that data availability and volume significantly impact the robustness of these metrics. Therefore, future users must carefully inspect data disparities and be cautious when interpreting results from regions with limited genome data.

Former Bayesian phylodynamic analyses, such as those conducted in Washington State ^47,48^, investigated the role of viral introductions in community spread. These studies use effective population sizes estimated from approximate structured coalescent models to determine the percentage of new cases resulting from introductions. Inspired by these studies, we propose integrating the Source Sink Score and Local Import Score into a Bayesian phylodynamic framework as future direction. This integration would allow us to calculate Bayesian Credible Intervals for these scores, providing a reliable measure of their uncertainty. This approach is particularly valuable when testing whether the Source Sink Score in one region, such as region A, is significantly higher than in another, such as region B, thereby facilitating robust regional comparisons.

## Author Contributions

Leke Lyu and Justin Bahl conceptualized and designed research. Leke Lyu, Gabriella Veytsel, Guppy Stott, Spencer Fox, Cody Dailey, Lambodhar Damodaran and Kayo Fujimoto performed research. Pamela Brown, Roger Sealy, Armand Brown and Magdy Alabady contributed new data. Leke Lyu, Gabriella Veytsel and Guppy Stott Analyzed data. Leke Lyu, Gabriella Veytsel, Guppy Stott, Spencer Fox, Cody Dailey, Lambodhar Damodaran, Kayo Fujimoto and Justin Bahl wrote and reviewd the paper. Justin Bahl acquired funding, supervised work, and coordinated communication among team members.

## Classification

Biological sciences / Ecology

## Data availability

The GISAID accession IDs of the genomes used in this study are provided on our GitHub repository (https://github.com/leke-lyu/transmissionCount). Additional data obtained during the study is available from the corresponding author upon reasonable request.

## Code availability

The R package *Subsamplerr*, which enables visual exploration of sampling heterogeneity and the implementation of proportional sampling schemes, is publicly available at https://github.com/leke-lyu/subsamplerr. For the pipeline setup and configurations used in the Nextstrain build, including Snakemake profiles, visit our GitHub repository at https://github.com/leke-lyu/surveillanceInTexas. All scripts used to generate the results in the Texas case study are publicly available at https://github.com/leke-lyu/transmissionCount.

## Data Availability

All data produced in the present study are available upon reasonable request to the authors

## Acknowledgments

This work has been funded in part from the National Institute of Allergy and Infectious Diseases, a component of the NIH, Department of Health and Human Services, under contract no. 75N93021C00018 (NIAID Centers of Excellence for Influenza Research and Response, CEIRR) and Centers for Disease Control and Prevention, Department of Health and Human Services, under contracts 75D30121C10133 and NU50CK000626. We acknowledge the GISAID contributors (acknowledgment table of genomes used is provided on our GitHub repository) for sharing genomic data.

