## Supplement Information for "Characterizing Spatial Epidemiology in a Heterogeneous Transmission Landscape Using a Novel Spatial Transmission Count Statistic"

**This PDF file includes:**

Genome dataset and subsampling scheme

Figure S1

Tables S1 to S3

Characterizing spatial transmission heterogeneity in subregions of Texas

Figures S2 to S26

Tables S4 to S5

Sensitivity Analysis

Table S6

Genome dataset and subsampling scheme

In Figure S1, we illustrate how our subsampling scheme reduces the sampling bias. Specifically: Figure S1A depicts the sampling ratio of the unsampled dataset, categorized by metropolitan areas and segmented by Epi-Week; Figure S1B shows the sampling ratio of the sampled dataset, also broken down by metropolitan areas.

In Table S1, we provide the complete list of metropolitan areas with their population and RUCC assignment. The RUCC form a classification scheme that distinguishes metropolitan counties by the population size of their metro area. The latest update is RUCC 2013, based on the 2010 United States census. Here, we updated the RUCC classification using the 2020 United States census. Only one change occurred: Bryan-College Station moved from RUCC-3 to RUCC-2 due to a population increase from 228,660 to 268,248.

In Table S2, we record the total case count, number of available genomes, and the number of sampled genomes from different subregions in Texas. The recorded time spans from Epi-Week 14 to Epi-Week 43.

In Table S3, we record the genome counts by country in the global contextual dataset. The complete list of GISAID’s accession IDs for SARS-CoV-2 sequences analyzed in this study is publicly available at <https://github.com/leke-lyu/transmissionCount>.

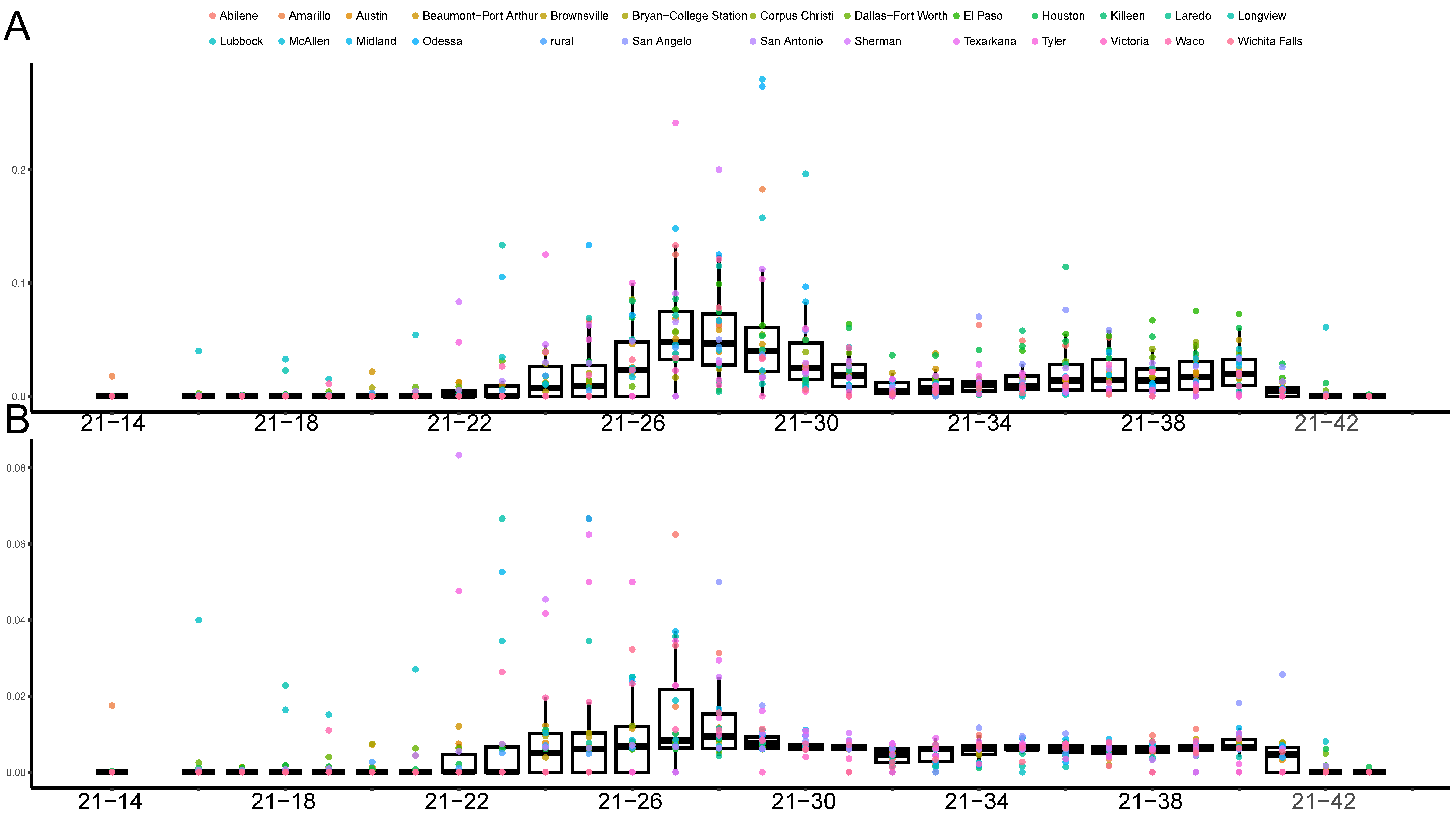

Figure S1. Genome sampling ratio of the original Texas dataset vs the sampled Texas dataset

Table S1. Population size of metropolitan areas in Texas and their corresponding RUCC

| **MAs** | **Population 2010** | **RUCC 2013** | **Population 2020** | **Updated RUCC** |
| --- | --- | --- | --- | --- |
| Dallas-Fort Worth | 6987215 | RUCC-1 | 7648780 | RUCC-1 |
| Houston | 5920416 | RUCC-1 | 7121540 | RUCC-1 |
| San Antonio | 2141508 | RUCC-1 | 2558143 | RUCC-1 |
| Austin | 1706289 | RUCC-1 | 2283371 | RUCC-1 |
| McAllen | 774769 | RUCC-2 | 870781 | RUCC-2 |
| El Paso | 625739 | RUCC-2 | 868859 | RUCC-2 |
| Killeen | 405300 | RUCC-2 | 475367 | RUCC-2 |
| Corpus Christi | 428185 | RUCC-2 | 445763 | RUCC-2 |
| Brownsville | 406220 | RUCC-2 | 421017 | RUCC-2 |
| Beaumont-Port Arthur | 403190 | RUCC-2 | 409782 | RUCC-2 |
| Lubbock | 290805 | RUCC-2 | 321368 | RUCC-2 |
| Waco | 252772 | RUCC-2 | 277547 | RUCC-2 |
| Amarillo | 251933 | RUCC-2 | 268691 | RUCC-2 |
| Bryan-College Station | 228660 | RUCC-3 | 268248 | RUCC-2 |
| Laredo | 250304 | RUCC-2 | 250304 | RUCC-2 |
| Tyler | 209714 | RUCC-3 | 233479 | RUCC-3 |
| Longview | 214369 | RUCC-3 | 217345 | RUCC-3 |
| Abilene | 165252 | RUCC-3 | 176579 | RUCC-3 |
| Midland | 141671 | RUCC-3 | 175220 | RUCC-3 |
| Odessa | 137130 | RUCC-3 | 165171 | RUCC-3 |
| Wichita Falls | 148415 | RUCC-3 | 148128 | RUCC-3 |
| Sherman | 120877 | RUCC-3 | 135543 | RUCC-3 |
| San Angelo | 111823 | RUCC-3 | 121516 | RUCC-3 |
| Victoria | 94003 | RUCC-3 | 98331 | RUCC-3 |
| Texarkana | 92565 | RUCC-3 | 92893 | RUCC-3 |

Table S2. Genomic sampling ratio by subregions in the Texas dataset

| **Reported Cases** | | **Available genomes (var = 5.74e-05)** | | **Sampled genomes (var = 7.56e-07)** | |
| --- | --- | --- | --- | --- | --- |
| Regions | Case Count | Genome Count | Ratio | Genome Count | Ratio |
| Dallas-Fort Worth | 331624 | 6621 | 0.020 | 1835 | 0.006 |
| Houston | 284582 | 10724 | 0.038 | 1523 | 0.005 |
| San Antonio | 108685 | 1922 | 0.018 | 593 | 0.005 |
| rural | 101957 | 958 | 0.009 | 516 | 0.005 |
| Austin | 88138 | 1329 | 0.015 | 465 | 0.005 |
| McAllen | 24147 | 245 | 0.010 | 113 | 0.005 |
| Killeen | 19473 | 393 | 0.020 | 78 | 0.004 |
| Corpus Christi | 17951 | 328 | 0.018 | 102 | 0.006 |
| Beaumont-Port Arthur | 14444 | 206 | 0.014 | 84 | 0.006 |
| Bryan-College Station | 13979 | 100 | 0.007 | 60 | 0.004 |
| El Paso | 13918 | 328 | 0.024 | 57 | 0.004 |
| Brownsville | 12961 | 222 | 0.017 | 66 | 0.005 |
| Lubbock | 10247 | 244 | 0.024 | 48 | 0.005 |
| Laredo | 8451 | 73 | 0.009 | 32 | 0.004 |
| Waco | 8045 | 113 | 0.014 | 48 | 0.006 |
| Amarillo | 7526 | 124 | 0.016 | 38 | 0.005 |
| Tyler | 7196 | 138 | 0.019 | 43 | 0.006 |
| Wichita Falls | 5972 | 41 | 0.007 | 26 | 0.004 |
| Victoria | 5502 | 29 | 0.005 | 21 | 0.004 |
| Longview | 5407 | 40 | 0.007 | 23 | 0.004 |
| Odessa | 4789 | 86 | 0.018 | 24 | 0.005 |
| Midland | 4436 | 100 | 0.023 | 29 | 0.007 |
| Sherman | 4315 | 58 | 0.013 | 25 | 0.006 |
| Abilene | 3487 | 87 | 0.025 | 22 | 0.006 |
| Texarkana | 2453 | 30 | 0.012 | 15 | 0.006 |
| San Angelo | 1870 | 54 | 0.029 | 13 | 0.007 |

Table S3. Genome counts by country in the global contextual dataset

| **Europe** | | **Asia** | | **North America** | | **South America** | | **Oceania** | | **Africa** | |
| --- | --- | --- | --- | --- | --- | --- | --- | --- | --- | --- | --- |
| United Kingdom | 1849 | Japan | 265 | USA | 2231 | Brazil | 78 | Australia | 41 | South Africa | 18 |
| Germany | 265 | India | 111 | Canada | 200 | Chile | 13 | New Zealand | 2 | Nigeria | 4 |
| Denmark | 214 | Indonesia | 14 | Mexico | 47 | Peru | 7 |  |  |  |  |
| France | 177 | Malaysia | 12 | Sint Maarten | 2 | Aruba | 2 |  |  |  |  |
| Sweden | 93 | Thailand | 9 |  |  |  |  |  |  |  |  |
| Switzerland | 83 | Philippines | 7 |  |  |  |  |  |  |  |  |
| Netherlands | 79 | Qatar | 3 |  |  |  |  |  |  |  |  |
| Italy | 78 | Vietnam | 3 |  |  |  |  |  |  |  |  |
| Spain | 78 | Bangladesh | 1 |  |  |  |  |  |  |  |  |
| Belgium | 63 | China | 1 |  |  |  |  |  |  |  |  |
| Ireland | 58 | Israel | 1 |  |  |  |  |  |  |  |  |
| Norway | 43 |  |  |  |  |  |  |  |  |  |  |
| Austria | 25 |  |  |  |  |  |  |  |  |  |  |
| Slovenia | 19 |  |  |  |  |  |  |  |  |  |  |
| Finland | 18 |  |  |  |  |  |  |  |  |  |  |
| Poland | 17 |  |  |  |  |  |  |  |  |  |  |
| Slovakia | 17 |  |  |  |  |  |  |  |  |  |  |
| Bulgaria | 15 |  |  |  |  |  |  |  |  |  |  |
| Croatia | 14 |  |  |  |  |  |  |  |  |  |  |
| Czech Republic | 14 |  |  |  |  |  |  |  |  |  |  |
| Lithuania | 13 |  |  |  |  |  |  |  |  |  |  |
| Estonia | 12 |  |  |  |  |  |  |  |  |  |  |
| Iceland | 10 |  |  |  |  |  |  |  |  |  |  |
| Russia | 10 |  |  |  |  |  |  |  |  |  |  |
| Romania | 8 |  |  |  |  |  |  |  |  |  |  |
| Greece | 6 |  |  |  |  |  |  |  |  |  |  |

Characterizing spatial transmission heterogeneity in subregions of Texas

In Figure S2, we present the time-adjusted phylogeny with branches colored based on ancestral trait reconstruction. The nextstrain build is available in: https://nextstrain.org/community/leke-lyu/deltaoutbreak/texas.

In Figure S3-S26, we summarized the epidemic trends in these subregions by utilizing the spatial transmission counts. The layout for the figures is as follows: the first row shows epidemic trends, the second row displays the Local Import Score, and the third row presents the Source Sink Score. Green lines represent importation, blue lines indicate local transmission, and pink lines signify exportation.

In Table S4, we present the complete centrality analysis of metropolitan areas in Texas using betweenness, connectedness, and degree measures.

In Table S5, we provide the complete set of Source Sink Scores and Local Import Scores for all subregions in Texas.

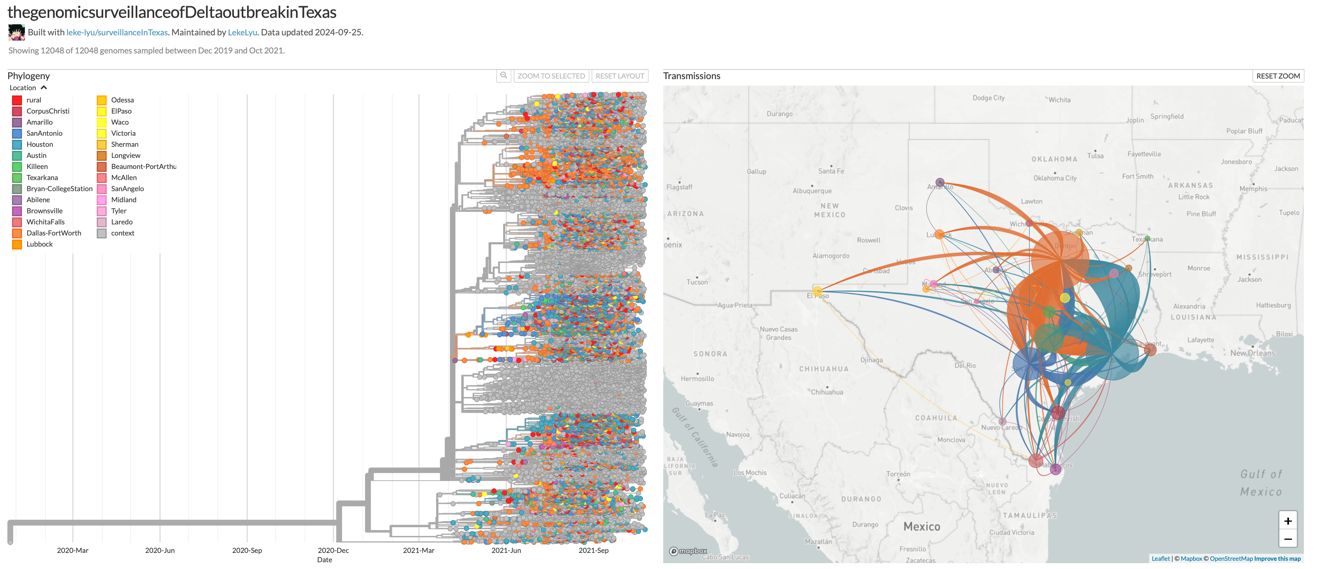

Figure S2. The time-adjusted phylogeny with branches colored based on ancestral trait reconstruction

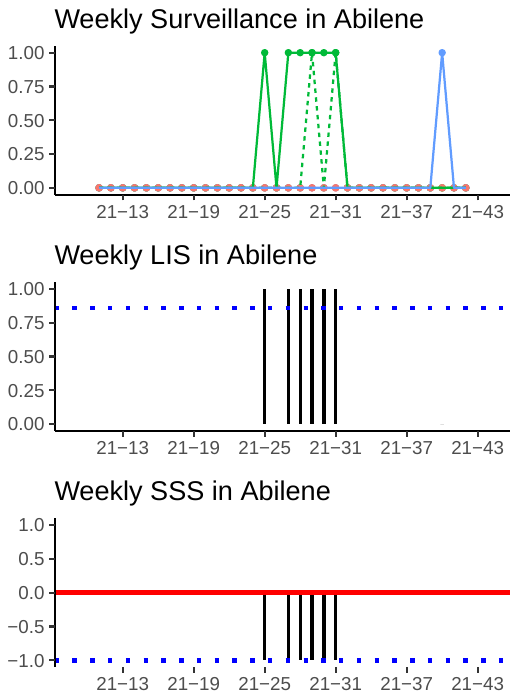

Figure S3. The epidemic trend in Abilence

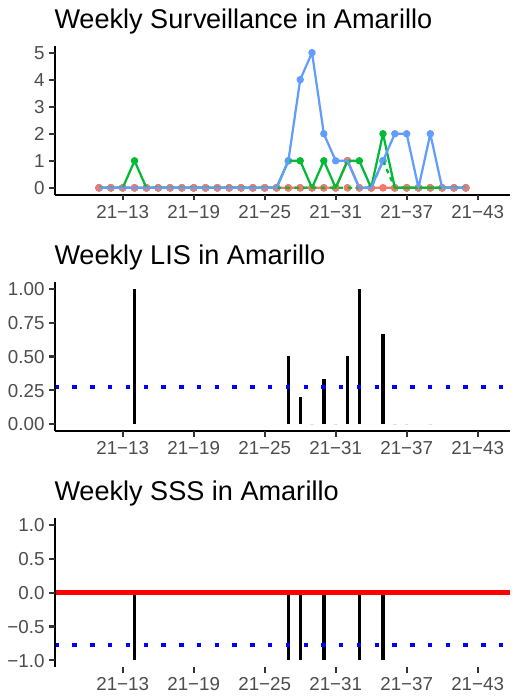

Figure S4. The epidemic trend in Amarillo

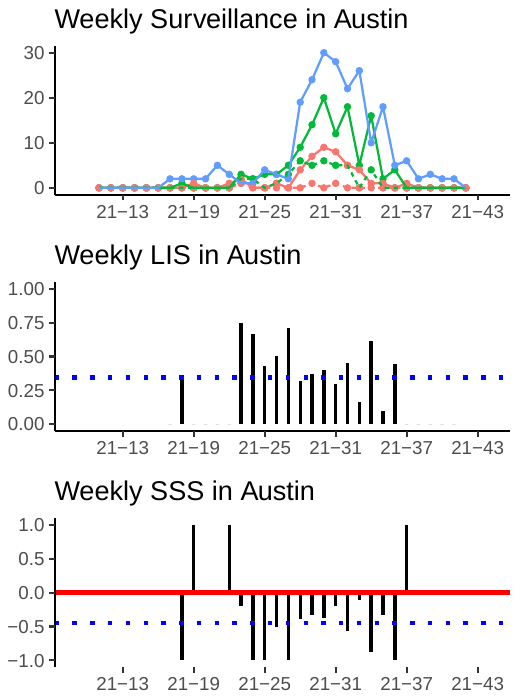

Figure S5. The epidemic trend in Austin

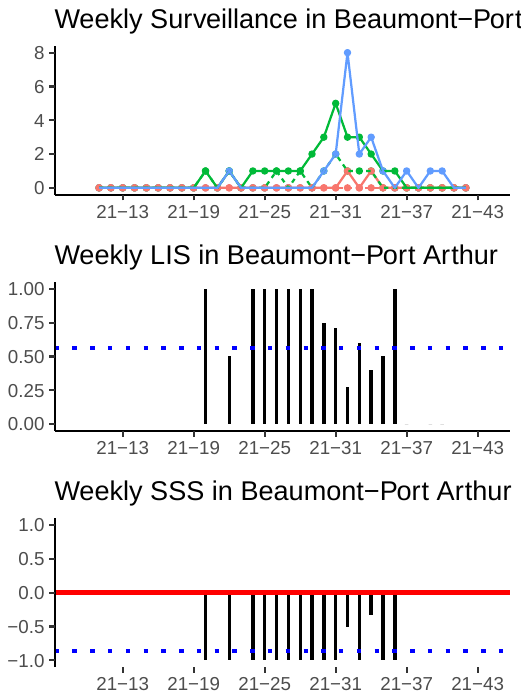

Figure S6. The epidemic trend in Beaumont-Port Arthur

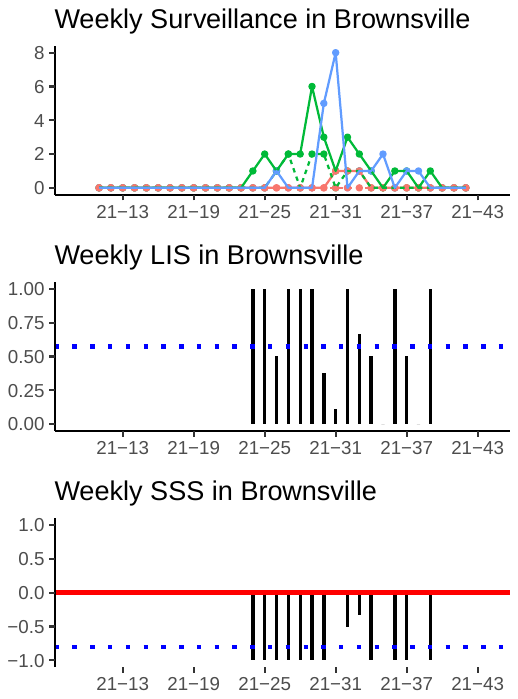

Figure S7. The epidemic trend in Brownsville

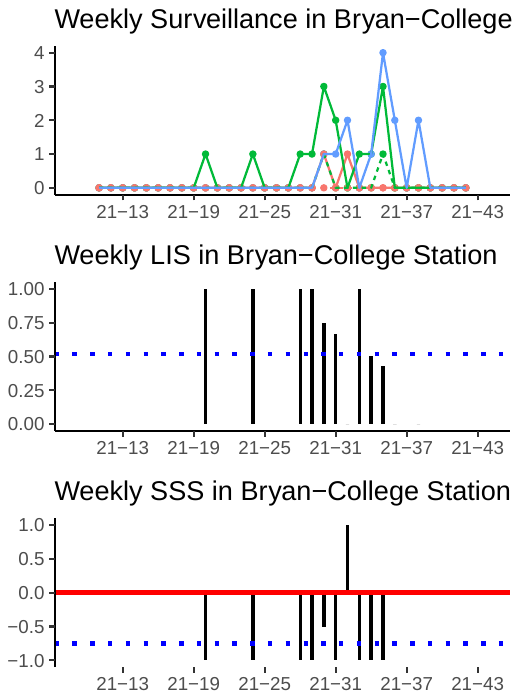

Figure S8. The epidemic trend in Bryan-College Station

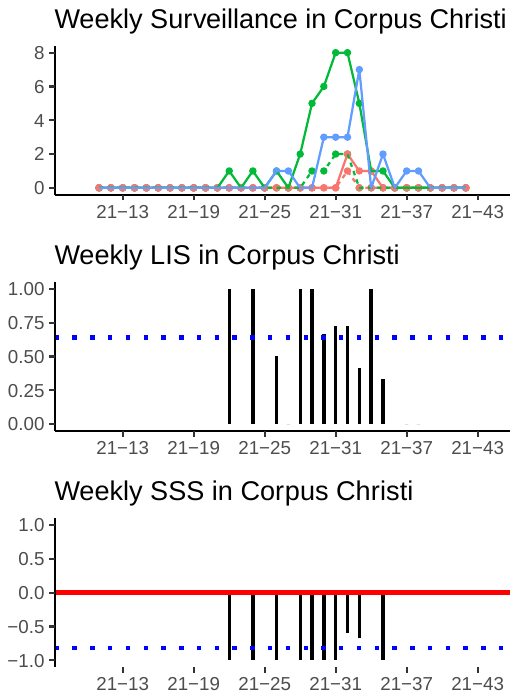

Figure S9. The epidemic trend in Corpus Christi

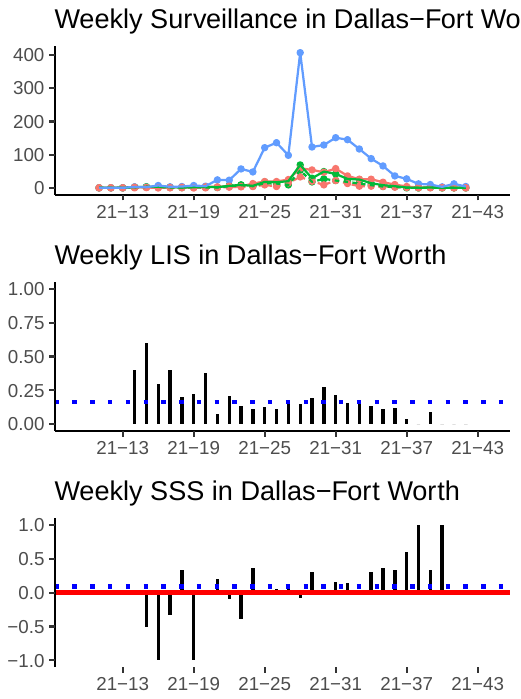

Figure S10. The epidemic trend in Dallas-Fort Worth

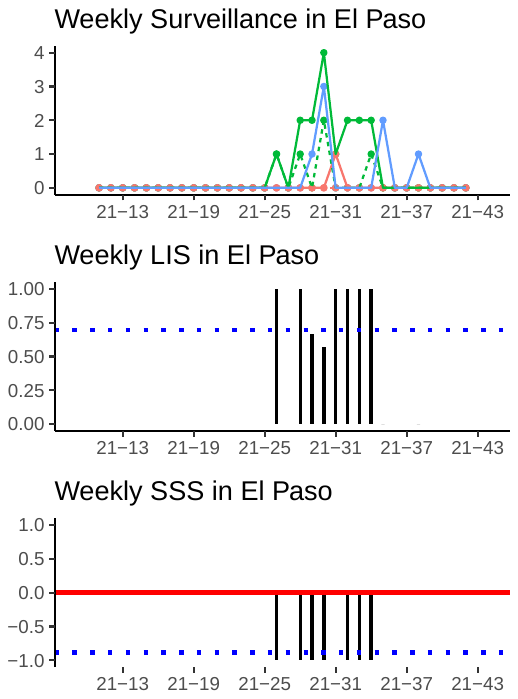

Figure S11. The epidemic trend in El Paso

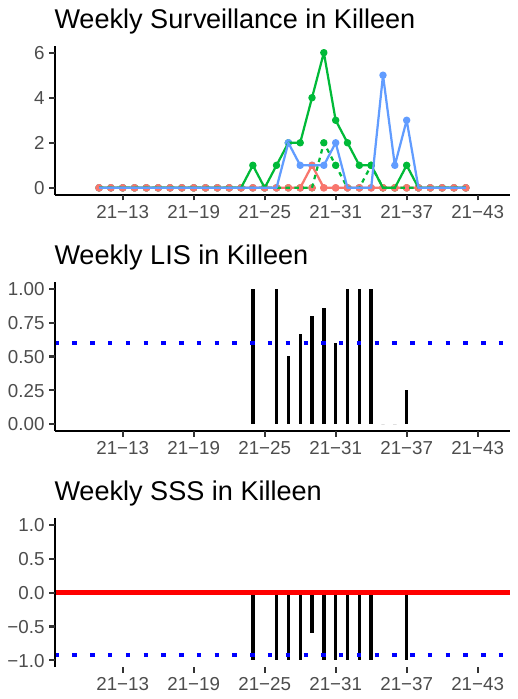

Figure S12. The epidemic trend in Killeen

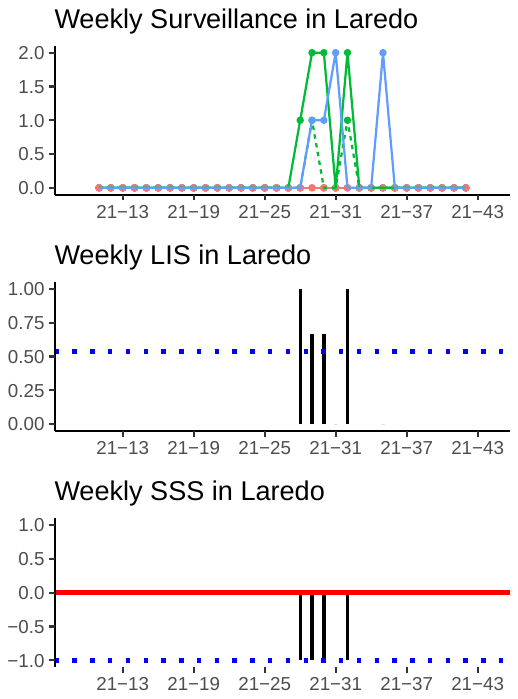

Figure S13. The epidemic trend in Laredo

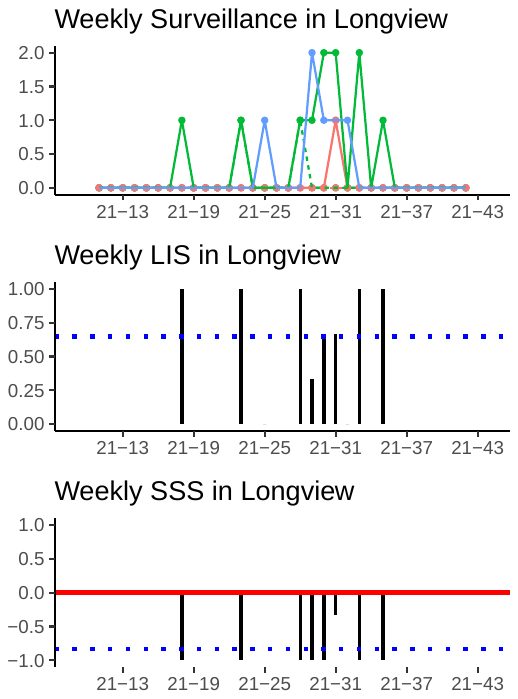

Figure S14. The epidemic trend in Longview

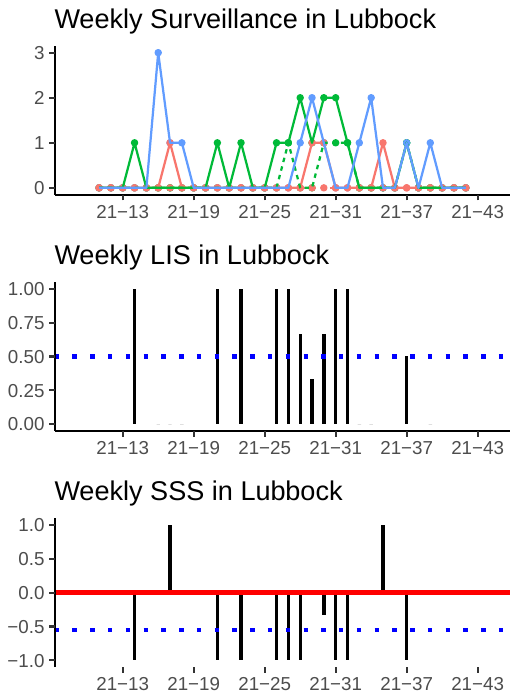

Figure S15. The epidemic trend in Lubbock

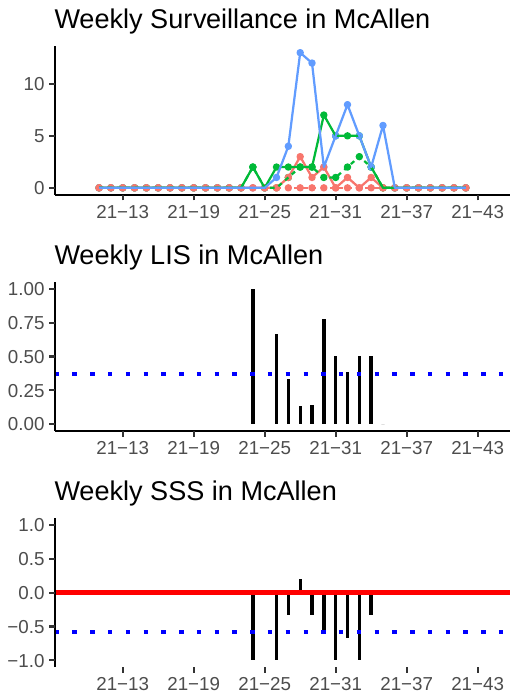

Figure S16. The epidemic trend in McAllen

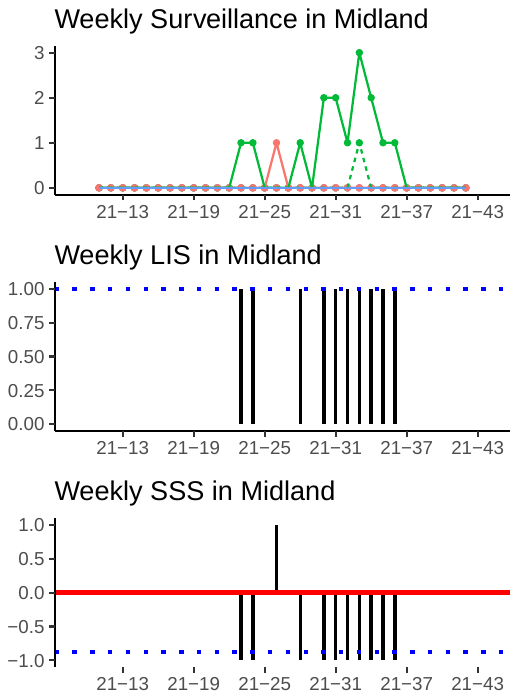

Figure S17. The epidemic trend in Midland

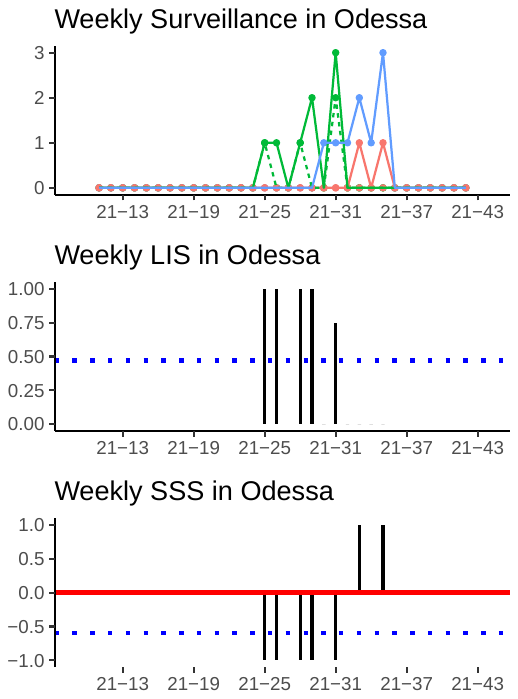

Figure S18. The epidemic trend in Odessa

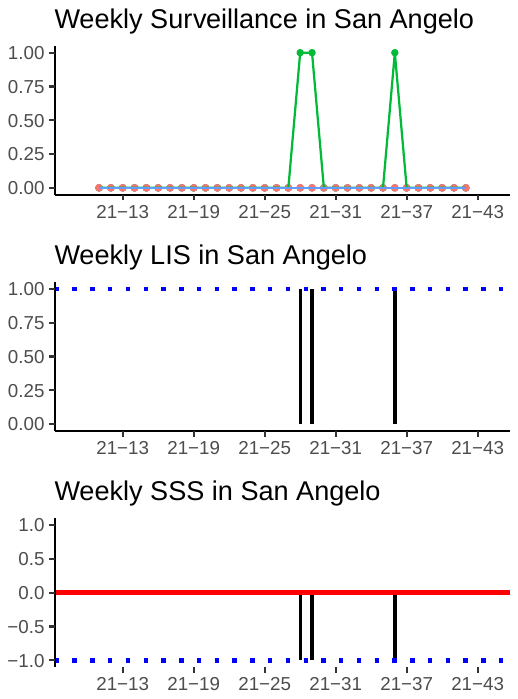

Figure S19. The epidemic trend in San Angelo

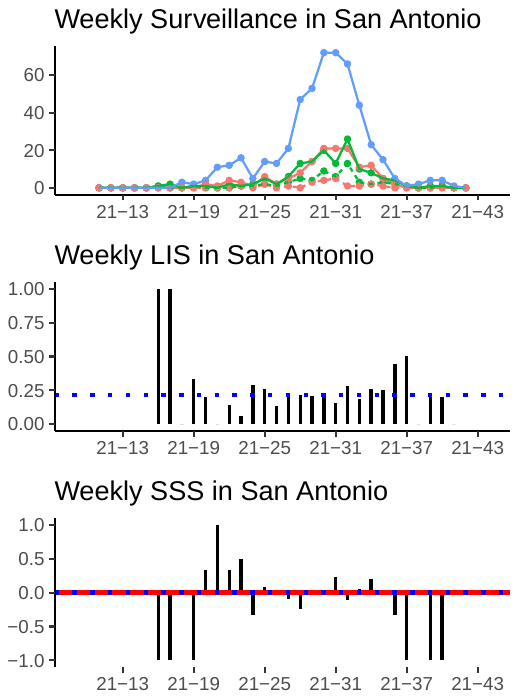

Figure S20. The epidemic trend in San Antonio

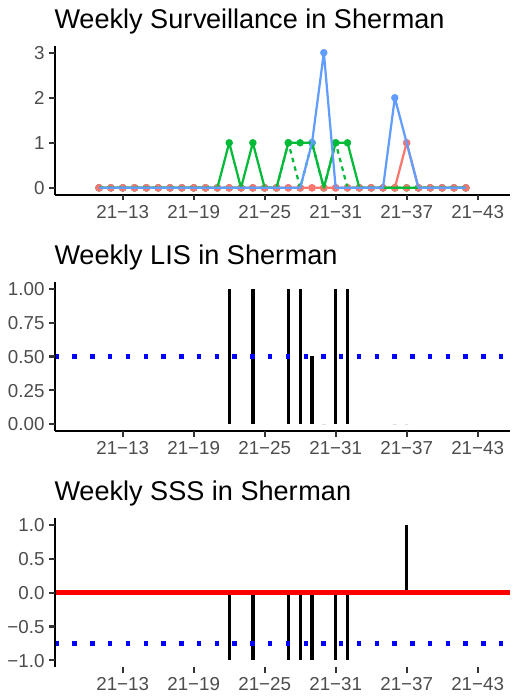

Figure S21. The epidemic trend in Sherman

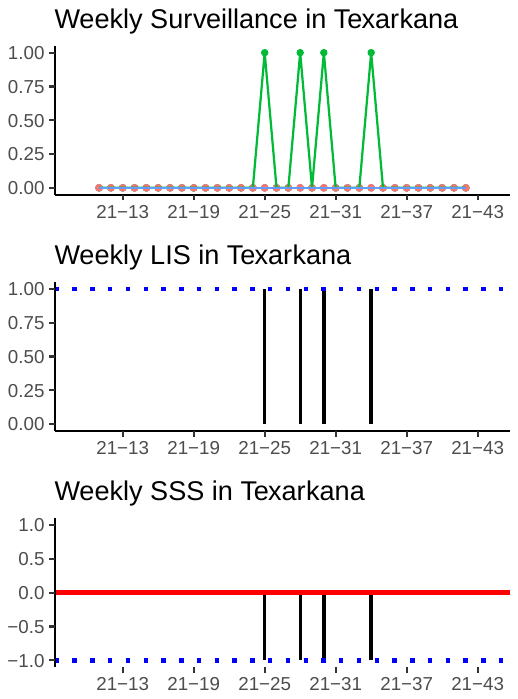

Figure S22. The epidemic trend in Texarkana

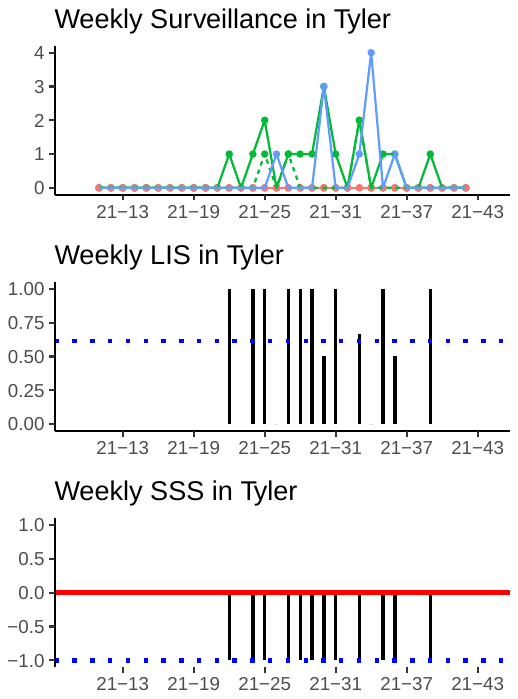

Figure S23. The epidemic trend in Tyler

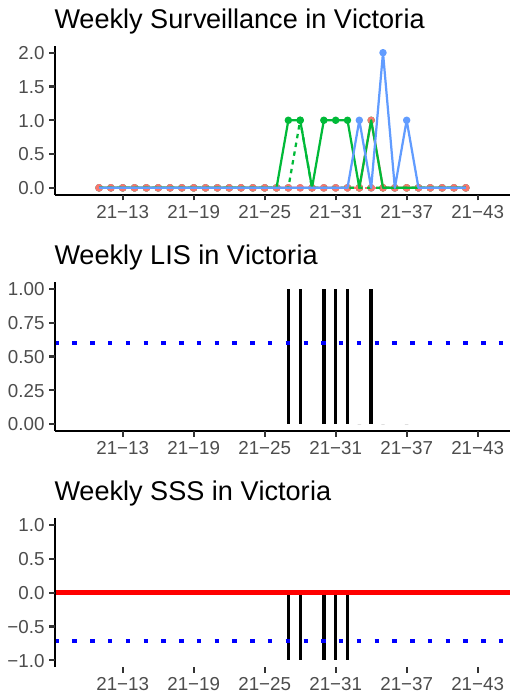

Figure S24. The epidemic trend in Victoria

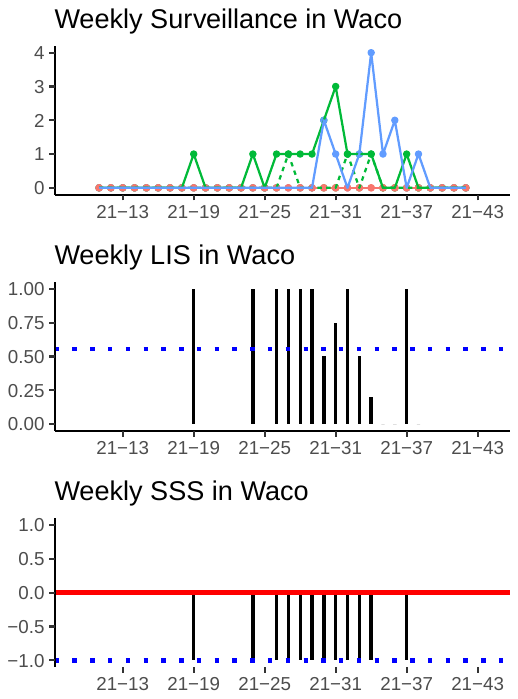

Figure S25. The epidemic trend in Waco

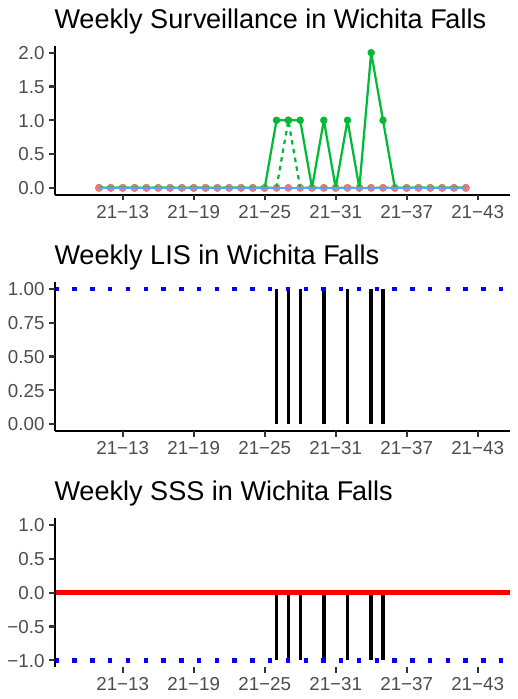

Figure S26. The epidemic trend in Wichita Falls

Table S4. Centrality analysis of metropolitan areas in Texas

| Metro areas | Betweenness | Closeness | Degree |
| --- | --- | --- | --- |
| Dallas-Fort Worth | 79.417 | 0.042 | 24 |
| Houston | 59.000 | 0.038 | 22 |
| San Antonio | 51.417 | 0.037 | 21 |
| Austin | 12.800 | 0.029 | 13 |
| McAllen | 3.200 | 0.026 | 9 |
| Midland | 1.200 | 0.024 | 7 |
| Bryan-College Station | 1.000 | 0.024 | 6 |
| Odessa | 0.667 | 0.023 | 5 |
| Corpus Christi | 0.600 | 0.024 | 7 |
| San Angelo | 0.500 | 0.023 | 5 |
| Brownsville | 0.500 | 0.024 | 6 |
| Lubbock | 0.250 | 0.023 | 5 |
| Abilene | 0.250 | 0.023 | 5 |
| Waco | 0.200 | 0.023 | 4 |
| Texarkana | 0.000 | 0.022 | 3 |
| Wichita Falls | 0.000 | 0.022 | 3 |
| Victoria | 0.000 | 0.022 | 4 |
| Tyler | 0.000 | 0.022 | 4 |
| Sherman | 0.000 | 0.022 | 3 |
| El Paso | 0.000 | 0.023 | 4 |
| Killeen | 0.000 | 0.022 | 4 |
| Amarillo | 0.000 | 0.022 | 4 |
| Laredo | 0.000 | 0.022 | 4 |
| Beaumont-Port Arthur | 0.000 | 0.022 | 3 |
| Longview | 0.000 | 0.022 | 3 |

Table S5. Source Sink Score and Local Import Score

| Metro areas | RUCC | SSS | LIS |
| --- | --- | --- | --- |
| Dallas-Fort Worth | 1 | 0.092 | 0.163 |
| Houston | 1 | 0.063 | 0.176 |
| San Antonio | 1 | 0.000 | 0.214 |
| Austin | 1 | -0.444 | 0.343 |
| Lubbock | 2 | -0.556 | 0.500 |
| McAllen | 2 | -0.581 | 0.370 |
| Odessa | 3 | -0.600 | 0.471 |
| Victoria | 3 | -0.714 | 0.600 |
| Sherman | 3 | -0.750 | 0.500 |
| Bryan-College Station | 2 | -0.750 | 0.519 |
| Amarillo | 2 | -0.778 | 0.276 |
| Brownsville | 2 | -0.800 | 0.574 |
| Corpus Christi | 2 | -0.814 | 0.639 |
| Longview | 3 | -0.833 | 0.647 |
| Beaumont-Port Arthur | 2 | -0.862 | 0.563 |
| Midland | 3 | -0.875 | 1.000 |
| El Paso | 2 | -0.882 | 0.696 |
| Killeen | 2 | -0.920 | 0.600 |
| Wichita Falls | 3 | -1.000 | 1.000 |
| Waco | 2 | -1.000 | 0.556 |
| Tyler | 3 | -1.000 | 0.615 |
| Texarkana | 3 | -1.000 | 1.000 |
| San Angelo | 3 | -1.000 | 1.000 |
| Laredo | 2 | -1.000 | 0.538 |
| Abilene | 3 | -1.000 | 0.857 |
| rural | 4-8 | -0.717 | 0.558 |

Sensitivity Analysis

Table S6 provides a detailed summary of the sensitivity analysis across all 10 replicates, which assess the robustness of the Source Sink Score and Local Import Score estimates.

Table S6. Sensitivity analysis

|  | Houston | Dallas-Fort Worth | San Antonio | Austin | McAllen | Odessa | rural | Corpus Christi | Amarillo | Longview | Lubbock | Beaumont-Port Arthur | Brownsville | Bryan-College Station | Midland | Killeen | Waco | Sherman | San Angelo | Wichita Falls | Tyler | Texarkana | Laredo | El Paso | Victoria | Abilene |
| --- | --- | --- | --- | --- | --- | --- | --- | --- | --- | --- | --- | --- | --- | --- | --- | --- | --- | --- | --- | --- | --- | --- | --- | --- | --- | --- |
| **SSS** | | | | | | | | | | | | | | | | | | | | | | | | | | |
| REP_0 | 0.190 | 0.084 | -0.325 | -0.458 | -0.375 | -0.500 | -0.624 | -0.610 | -0.846 | -0.818 | -0.889 | -0.867 | -0.813 | -0.917 | -1.000 | -0.933 | -0.750 | -1.000 | -1.000 | -1.000 | -0.733 | -0.667 | -1.000 | -1.000 | -1.000 | -0.750 |
| REP_1 | 0.092 | 0.073 | -0.071 | -0.390 | -0.464 | -0.800 | -0.621 | -0.636 | -0.400 | -0.875 | -0.556 | -0.800 | -0.615 | -0.923 | -0.500 | -0.871 | -1.000 | -1.000 | -1.000 | -1.000 | -1.000 | -1.000 | -1.000 | -1.000 | -1.000 | -1.000 |
| REP_2 | 0.118 | 0.150 | -0.100 | -0.449 | -0.440 | -0.200 | -0.727 | -0.800 | -0.455 | -1.000 | -1.000 | -0.765 | -0.935 | -0.800 | -1.000 | -0.852 | -1.000 | -1.000 | -1.000 | -1.000 | -1.000 | -1.000 | -0.833 | -1.000 | -1.000 | -1.000 |
| REP_3 | 0.087 | 0.115 | -0.081 | -0.516 | -0.333 | -0.667 | -0.619 | -0.581 | -0.778 | -0.714 | -1.000 | -0.813 | -0.742 | -0.800 | -1.000 | -1.000 | -0.895 | -0.778 | -1.000 | -1.000 | -1.000 | -1.000 | -1.000 | -0.909 | -1.000 | -1.000 |
| REP_4 | 0.158 | 0.049 | -0.015 | -0.513 | -0.659 | -0.500 | -0.735 | -0.750 | -0.143 | -0.571 | -0.750 | -0.935 | -0.926 | -0.818 | -0.867 | -0.778 | -0.818 | -1.000 | -1.000 | -1.000 | -1.000 | -1.000 | -1.000 | -1.000 | -1.000 | -1.000 |
| REP_5 | 0.077 | 0.127 | -0.135 | -0.351 | -0.302 | -0.500 | -0.673 | -0.739 | -0.833 | -0.818 | -0.545 | -0.867 | -1.000 | -0.481 | -0.846 | -0.947 | -0.909 | -1.000 | -1.000 | -1.000 | -1.000 | -1.000 | -1.000 | -1.000 | -1.000 | -1.000 |
| REP_6 | 0.063 | 0.092 | 0.000 | -0.444 | -0.581 | -0.600 | -0.717 | -0.814 | -0.778 | -0.833 | -0.556 | -0.862 | -0.800 | -0.750 | -0.875 | -0.920 | -1.000 | -0.750 | -1.000 | -1.000 | -1.000 | -1.000 | -1.000 | -0.882 | -0.714 | -1.000 |
| REP_7 | 0.146 | 0.083 | -0.198 | -0.466 | -0.583 | -0.500 | -0.617 | -0.733 | -0.714 | -0.692 | -0.905 | -0.771 | -0.667 | -0.909 | -0.571 | -0.933 | -0.778 | -0.818 | -0.333 | -1.000 | -1.000 | -1.000 | -0.857 | -1.000 | -1.000 | -1.000 |
| REP_8 | 0.127 | 0.108 | 0.054 | -0.440 | -0.882 | -1.000 | -0.573 | -0.551 | -0.800 | -0.714 | -1.000 | -0.784 | -0.813 | -0.909 | -1.000 | -0.867 | -0.810 | -1.000 | -1.000 | -0.778 | -0.714 | -1.000 | -1.000 | -1.000 | -1.000 | -1.000 |
| REP_9 | 0.174 | 0.152 | -0.208 | -0.583 | -0.633 | -0.333 | -0.554 | -0.321 | -0.833 | -0.692 | -0.739 | -0.688 | -0.889 | -0.895 | -1.000 | -0.733 | -1.000 | -0.778 | -1.000 | -0.600 | -1.000 | -1.000 | -1.000 | -0.917 | -1.000 | -1.000 |
| **LIS** | | | | | | | | | | | | | | | | | | | | | | | | | | |
| REP_0 | 0.153 | 0.157 | 0.275 | 0.385 | 0.367 | 0.529 | 0.515 | 0.440 | 0.444 | 0.588 | 0.810 | 0.528 | 0.644 | 0.719 | 0.750 | 0.592 | 0.600 | 0.667 | 0.800 | 0.889 | 0.591 | 0.625 | 0.769 | 0.846 | 0.625 | 1.000 |
| REP_1 | 0.165 | 0.160 | 0.220 | 0.353 | 0.466 | 0.600 | 0.538 | 0.450 | 0.200 | 0.833 | 0.636 | 0.443 | 0.583 | 0.714 | 0.545 | 0.690 | 0.739 | 1.000 | 1.000 | 0.909 | 0.636 | 0.571 | 0.722 | 0.696 | 1.000 | 0.571 |
| REP_2 | 0.171 | 0.153 | 0.214 | 0.353 | 0.375 | 0.500 | 0.518 | 0.529 | 0.276 | 0.938 | 0.826 | 0.566 | 0.732 | 0.563 | 0.929 | 0.735 | 0.846 | 0.800 | 1.000 | 1.000 | 0.625 | 1.000 | 0.647 | 0.950 | 0.889 | 0.700 |
| REP_3 | 0.178 | 0.157 | 0.213 | 0.370 | 0.350 | 0.588 | 0.549 | 0.386 | 0.235 | 0.706 | 0.724 | 0.518 | 0.628 | 0.486 | 0.909 | 0.718 | 0.621 | 0.615 | 0.667 | 0.889 | 0.652 | 0.571 | 0.833 | 0.875 | 0.571 | 1.000 |
| REP_4 | 0.156 | 0.169 | 0.205 | 0.401 | 0.405 | 0.750 | 0.545 | 0.500 | 0.286 | 0.550 | 0.560 | 0.625 | 0.650 | 0.645 | 0.667 | 0.727 | 0.645 | 0.583 | 1.000 | 1.000 | 0.619 | 0.750 | 0.818 | 0.870 | 0.714 | 0.889 |
| REP_5 | 0.171 | 0.154 | 0.238 | 0.356 | 0.326 | 0.462 | 0.549 | 0.460 | 0.524 | 0.526 | 0.630 | 0.459 | 0.600 | 0.606 | 0.923 | 0.902 | 0.700 | 0.889 | 1.000 | 0.833 | 0.619 | 1.000 | 0.615 | 0.778 | 1.000 | 0.750 |
| REP_6 | 0.176 | 0.163 | 0.214 | 0.343 | 0.370 | 0.471 | 0.558 | 0.639 | 0.276 | 0.647 | 0.500 | 0.563 | 0.574 | 0.519 | 1.000 | 0.600 | 0.556 | 0.500 | 1.000 | 1.000 | 0.615 | 1.000 | 0.538 | 0.696 | 0.600 | 0.857 |
| REP_7 | 0.173 | 0.155 | 0.245 | 0.386 | 0.481 | 0.529 | 0.543 | 0.582 | 0.429 | 0.550 | 0.769 | 0.564 | 0.595 | 0.656 | 0.611 | 0.707 | 0.640 | 0.769 | 0.400 | 0.909 | 0.600 | 1.000 | 0.619 | 0.714 | 0.800 | 0.667 |
| REP_8 | 0.156 | 0.151 | 0.183 | 0.359 | 0.571 | 0.909 | 0.445 | 0.481 | 0.391 | 0.632 | 0.789 | 0.569 | 0.644 | 0.724 | 0.833 | 0.737 | 0.792 | 0.733 | 1.000 | 0.727 | 0.600 | 0.800 | 0.688 | 0.759 | 0.833 | 0.750 |
| REP_9 | 0.152 | 0.142 | 0.245 | 0.397 | 0.494 | 0.471 | 0.519 | 0.455 | 0.314 | 0.647 | 0.690 | 0.529 | 0.654 | 0.545 | 0.929 | 0.650 | 0.875 | 0.727 | 0.500 | 0.400 | 0.667 | 1.000 | 0.692 | 0.793 | 0.800 | 0.800 |
